# *Neuropeptide S* as a potential risk *locus* for migraine in the Portuguese population

**DOI:** 10.64898/2026.01.20.26344443

**Authors:** Rodrigo De Marco, Kevin Pucci, Mariana Santos, Raquel Gil-Gouveia, Bruno Cavadas, Alda Sousa, Miguel Alves-Ferreira, Luísa Azevedo, Carolina Lemos, Andreia Dias

## Abstract

**Objective:** The most common forms of migraine are complex disorders characterized by significant clinical diversity. The genetic basis of migraine has been the subject of many studies but remains largely unknown. We present the first pilot genome-wide association study (GWAS) integrating a polygenic risk score (PRS) in the Portuguese population, designed to identify migraine susceptibility risk *loci* through a case-control study, in order to unravel population-specific variants.

**Methods:** Genotyping was conducted on 380 individuals using the Applied Biosystems Axiom™ PMDA array. A vast dataset of 8,031,293 single-nucleotide polymorphisms (SNPs) was available, providing a comprehensive scope for GWAS analysis. PRS models were created and tested on subsets of the genotyping data, and the optimal statistical significance threshold was assessed.

**Results:** We detected nine risk *loci* corresponding to nine lead SNPs (*ZNF385D*, *YTHDF3*, *NPS*, *RAP1A/INKA2*, *CTA-481E9.4/CTA-481E9.3*, *AC092079.1*, *PPHLN1*, *SMYD3* and *AL355597.1* (near *ADARB2*)). Additionally, eleven variants, without any previous association to pathogenicity in migraine, were highlighted by the chosen PRS model. Among our results, a new *locus* within the *NPS* gene, representing a novel association with migraine, is a potential new target directly related to recent and effective migraine treatments.

**Conclusions:** These findings reinforce the importance of neurotransmitter release and synaptic transmission, as well as the involvement of vascular components, in migraine pathophysiology. This work underscores that GWAS can provide novel, clinically valuable insights into populational and disease-associated genetic landscapes, enabling therapeutic developments and bolstering personalized medicine strategies.

## INTRODUCTION

The genetics of migraine has been studied since the nineteenth century when Liveing and Tissot first described the heritability of migraine ^1^. The most common forms of migraine with and without aura (MwA/MwoA) are complex disorders characterized by significant clinical heterogeneity. Migraine was initially attributed to vascular mechanisms, but subsequent research has underscored the relevance of neuronal mechanisms ^2^.

The pathophysiology of these disorders is only partially understood, but involves the activation of the trigeminovascular system, responsible for pain sensation. Activation of these pathways mediates the release of neuropeptides, such as calcitonin gene-related peptide (CGRP), substance P (SP), neurokinin A, vasoactive intestinal peptide (VIP), pituitary adenylate cyclase-activating peptide (PACAP) and nitric oxide ^3,4^. Many of the migraine therapies act on CGRP release, including triptans, ditans, CGRP receptor blockers (gepants) and CGRP-related monoclonal antibodies, now considered a breakthrough in migraine-specific treatments ^5^. In addition to CGRP, the PACAP pathway might prove to be a useful neuropeptide system for preventing and/or blocking migraine attacks.

Genome-wide association studies (GWAS) in migraine have allowed the identification of several *loci* that harbor genetic risk factors ^6^, which have low penetrance individually, but together might have a significant impact on disease susceptibility. More recently, a GWAS meta-analysis of migraine reported 123 genomic *loci*, of which 86 were previously unknown, and included new risk *loci* containing target genes (*CALCA* and *CALCB*) associated with the CGRP pathway and the serotonin 5-HT1F receptor ^7^. Nonetheless, population-specific GWAS remain crucial to understand intrapopulation genetics’ heterogeneity ^8^.

Given the polygenic nature of migraine, several statistical methods have gained prominence in recent years, with the polygenic risk score (PRS) standing out as a valuable tool to quantify the relative risk of a given phenotype. It is computed as a linear combination of single nucleotide polymorphisms (SNPs), where each SNP is weighted by its effect size and then summed (*PRS* = Σ *SNPi* × *βi*, with *SNPi* representing the effect allele count and *βi* the effect size) ^9^. By accounting for multiple SNPs simultaneously, it is possible to increase the predictive power of the model ^10^.

Describing population-specific *loci* is crucial to unraveling the pathophysiology of migraine, so we present herein the first GWAS in the Portuguese population, aimed at identifying migraine susceptibility risk *loci* through a case-control study. We then generated a model to calculate the PRS, allowing the identification of the SNPs most strongly associated with migraine phenotypes.

## MATERIALS AND METHODS

### Subjects and Study Design

This case-control study was conducted in a Portuguese cohort from the outpatient neurologic clinic at Centro Hospitalar Universitário de Santo António (CHUdSA) and at Centre for Predictive and Preventive Genetics - Institute for Research and Innovation in Health (CGPP-i3S). We carried out a GWAS analysis of 380 individuals: 190 migraine patients and 190 controls. Clinical information of subjects was collected, and patients with FHM were excluded; those with the co-occurrence of MwA and MwoA were included in the MwA group. Controls and cases were from the same ethnic and geographical origin, age-matched and non-related. All cases and controls underwent a diagnostic interview, using the same structured questionnaire, based on the operational criteria of the International Headache Society (IHS)—3rd edition of the International Classification of Headache Disorders (ICHD-3) ^11^. Blood samples were collected in the sequence of a neurologic appointment and were stored at CGPP’s biobank at i3S. The Ethics Committees of CHUdSA and i3S approved the study and participants gave their written informed consent.

### DNA Extraction and Genome-Wide Array Genotyping

Genomic DNA extraction from peripheral blood samples was performed by the standard salting-out method using the QIAamp® DNA Blood Mini Kit ^12^. DNA quantification was performed using NanoDrop™ One. The genotyping of over 900,000 SNPs (900 K) was attained with the Axiom™ Precision Medicine Diversity Array (PMDA, Affymetrix) and the GeneTitan Multi-Channel (MC) Instrument (Thermo Fisher Scientific, Waltham, MA, USA). Genotyping raw data was analyzed with the Axiom™ Analysis Suite version 5.1 (Applied Biosystems), using the Best Practices Workflow with default settings.

### Imputation and Quality Control

Genome-wide data were imputed using the Haplotype Reference Consortium (HRC) panel (r1.1) on the Sanger Imputation Server ^13^ applying EAGLE2 as a pre-phasing method and Positional Burrows-Wheeler Transform (PBWT) for imputation ^14^.

Quality control (QC) of genotyped data was accessed and filtered for minor allele frequency (MAF) > 0.01, sample call rate > 0.95, marker call rate > 0.95 and Hardy-Weinberg equilibrium (HWE) *p*-value > 1.0×10^-6^. After imputation and QC, a total of 8,031,293 SNPs remained for the final analyzes.

### GWAS and statistical analysis

Only genotyped variants with *p*-values below genome-wide significance (*p* = 5 × 10^−8^) were considered. Manhattan plots and quantile-quantile (QQ) plots were generated using the FUMA software ^15^. The *locus* regional plots were obtained from the LocusZoom software ^16^.

### Identification of lead and candidate SNPs associated with migraine

An association analysis for each SNP with migraine susceptibility was performed using PLINK v1.90b6.26 ^17^, based on a χ^2^ test for each SNP to the phenotype of interest. Independent significant SNPs (r^2^ < 0.6) were filtered with FUMA software ^15^ with *p*-values below Bonferroni-corrected genome-wide significance (two-tailed *p* < 5 × 10^−8^). All SNPs in LD (r^2^ ≥ 0.6) to one of the independent significant SNPs were defined as candidate SNPs. All independent significant SNPs with r^2^ < 0.1 were defined as lead SNPs and merged into one *locus* if located within 250kb of one another. These LD analyses were conducted assuming the 1000G Phase3 EUR reference panel.

### Gene mapping

Functional annotations of SNPs were obtained from: CADD (12.37 as the threshold for a deleterious score), RegulomeDB (maximum score of 7) and 15-core chromatin states. Additionally, only significant expression quantitative trait *loci* (eQTL) values (false discovery rate (FDR) < 0.05), according to GTEx V8, were selected to map SNPs to genes.

Regulatory genomic data (such as enhancers and transcription factor (TF) binding sites) was analyzed to interpret the impact of non-coding variants ^18^. FUMA results were complemented by HaploReg v4.2 (r^2^ > 0.8) analysis, exploring variants in LD with lead SNPs.

### Enrichment analysis

To identify known biological pathways and gene sets at the associated *loci*, an enrichment approach, using EnrichR platform ^19,20^ was performed with the Gene Ontology (GO) ^21^, Kyoto Encyclopaedia of Genes and Genomes (KEGG) ^22^, and Reactome ^23^ databases.

### Protein interactions

STRING ^24^ (v12.0) was used to search for protein-protein interactions (PPI), including direct (physical) and indirect (functional) associations. The minimum required interaction score was set at 0.400.

### Polygenic risk score

For quality control and PRS calculation, we used the PLINK software ^17^, as previously described ^25^. The complete dataset was subdivided into a base dataset and a target dataset, consisting of 190 individuals each, divided into 95 cases and 95 controls. MAF was estimated exclusively from the base dataset.

Quality control was performed according to established PLINK guidelines, and confirming that no duplicate SNPs were detected in either the base or target dataset, ambiguous SNPs were not an issue due to identical generation conditions, and no samples overlapped between the base and target datasets.

SNPs were excluded based on low MAF (cut-off of 0.01), significant deviation from Hardy–Weinberg equilibrium (*p*-value < 10⁻⁶) and SNPs with data missing in a considerable fraction of subjects (cut-off of 0.01).

The PRS was calculated using the clumping and thresholding method implemented in PLINK. To account for linkage disequilibrium (LD), clumping was performed with the following parameters: *p*-value ≤ 1 (thus including all SNPs), r² ≥ 0.1, and clump all SNPs with distance inferior to 250kb. Thresholding was then applied to exclude SNPs with high *p*-values, reducing noise and potential overfitting. In line with the recommended guidelines ^25^, PRSs were computed across multiple significance thresholds (5×10⁻⁹, 1×10⁻⁸, 1×10⁻⁷, 1×10⁻⁶, 5×10⁻⁵, 1×10⁻⁴, 2×10⁻⁴, 5×10⁻⁴, 1×10⁻³, 5×10⁻², 0.1, 0.2, 0.3, 0.4, and 0.5) to identify the optimal predictive model, as the best threshold is not known *a priori*. Model performance was evaluated using McFadden’s pseudo-R², and results were visualized by plotting pseudo-R² against *p*-value thresholds and comparing PRS distributions between cases and controls in the validation data. Visualization of PRS results was performed using RStudio version 4.4.2 (2024-10-31 ucrt) ^26^.

## RESULTS

### Characterization of sample study

We studied 380 individuals, with a case-control ratio of 1:1. Concerning migraineurs, 95 were MwA and 95 were MwoA patients. At the time of observation, the mean age of controls was slightly higher than that of migraineurs (38.80 ± 14.52 vs. 35.01 ± 13.15 years), increasing confidence that controls are, in fact, migraine-free (**Table 1**). Given the limited sample size, no stratification of migraineurs into MwA or MwoA subgroups was performed.

**Table 1.** Demographic and clinical data of migraine patients and controls.

|  | Migraineurs (n=190) | Controls (n=190) |
| --- | --- | --- |
| Migraine with Aura (MA) | 95 | n/a |
| Migraine without Aura (MO) | 95 | n/a |
| Mean age at observation (±SD) years | 35.01 (±) 13.15 | 38.80 (±) 14.52 |

SD – standard deviation; MA – migraine with aura; MO – migraine without aura; n/a – not applicable.

### Identification and characterization of migraine risk *loci*

We found nine genomic risk *loci* corresponding to nine lead SNPs (six intronic, two intergenic and one ncRNA SNP). Regarding variants in high LD (r^2^ > 0.6) with the lead SNPs (candidate SNPs), we found 223 variants (**Supplementary Table 1**), of which 54 (24.2%) were located within protein-coding genes, 156 (70%) were intergenic, and 13 (5.8%) were ncRNA variants. Regarding the 54 protein-coding variants, two were exonic located in the *YTHDF3* and *NPS* genes. Two of the 223 candidate SNPs were considered deleterious according to the CADD-PHRED score (12.37) ^27^, suggesting a potential functional impact (rs6537713, in *RAP1A*/*INKA2* genes; rs4775219, in RP11-568G20.2) (**Supplementary Table 2**). Additionally, we identified six genome-wide significant *loci* with mapped genes (*ZNF385D*, *YTHDF3*, *NPS*, *RAP1A/INKA2*, *PPHLN1* and *SMYD3*). The other three risk *loci* exist in intergenic regions (CTA-481E9.4/CTA-481E9.3; AC092079.1; AL355597.1 (near *ADARB2*)) (**Table 2**).

**Table 2.** Characterization of 11 genomic risk *loci* of interest.

| Locus | SNP | CHR | Position (GrCh37) | A1 | A2 | MAF | CHISQ | P | OR |
| --- | --- | --- | --- | --- | --- | --- | --- | --- | --- |
| *ZNF385D* | rs9812687 | 3 | 22177068 | T | C | 0.2495 | 65.21 | $6.75 \times 10^{-16}$ | 3.350 |
| *YTHDF3* | rs4739066 | 8 | 64104092 | G | A | 0.0716 | 56.12 | $6.83 \times 10^{-14}$ | 3.753 |
| *NPS* | rs11018195 | 10 | 129349946 | G | A | 0.0477 | 52.62 | $4.04 \times 10^{-13}$ | 0.297 |
| *RAP1A*<br>*INKA2* | rs61791765 | 1 | 112223854 | C | A | 0.0785 | 40.68 | $1.79 \times 10^{-10}$ | 2.905 |
| CTA-481E9.4<br>CTA-481E9.3 | rs4781860 | 16 | 18187679 | G | A | 0.1471 | 39.81 | $2.79 \times 10^{-10}$ | 2.649 |
| AC092079.1 | rs9806148 | 15 | 60158604 | G | A | 0.0497 | 37.75 | $8.03 \times 10^{-10}$ | 2.877 |
| *PPHLN1* | rs117423705 | 12 | 42818514 | T | C | 0.0189 | 35.50 | $2.54 \times 10^{-9}$ | 2.768 |
| *SMYD3* | rs1934582 | 1 | 245975417 | C | T | 0.1928 | 33.63 | $6.68 \times 10^{-9}$ | 0.361 |
| AL355597.1<br>(near *ADARB2*) | rs118096186 | 10 | 1959505 | T | C | 0.0199 | 33.60 | $6.75 \times 10^{-9}$ | 2.875 |

CHR – chromosome; A1 – effect allele; A2 – non-effect allele; MAF – minor allele frequency; CHISQ – basic allelic test chi-square; P – *p*-value; OR – odds ratio.

A Manhattan plot was generated from the resulting data of 8,031,293 SNPs associated with migraine susceptibility (**Fig 1A**). The QQ plot (**Fig 1B**) and the genomic inflation factor (lambda, λ = 1.076 – indicating that the *p*-values were not inflated) showed no evidence of systematic bias.

**Figure 1:**
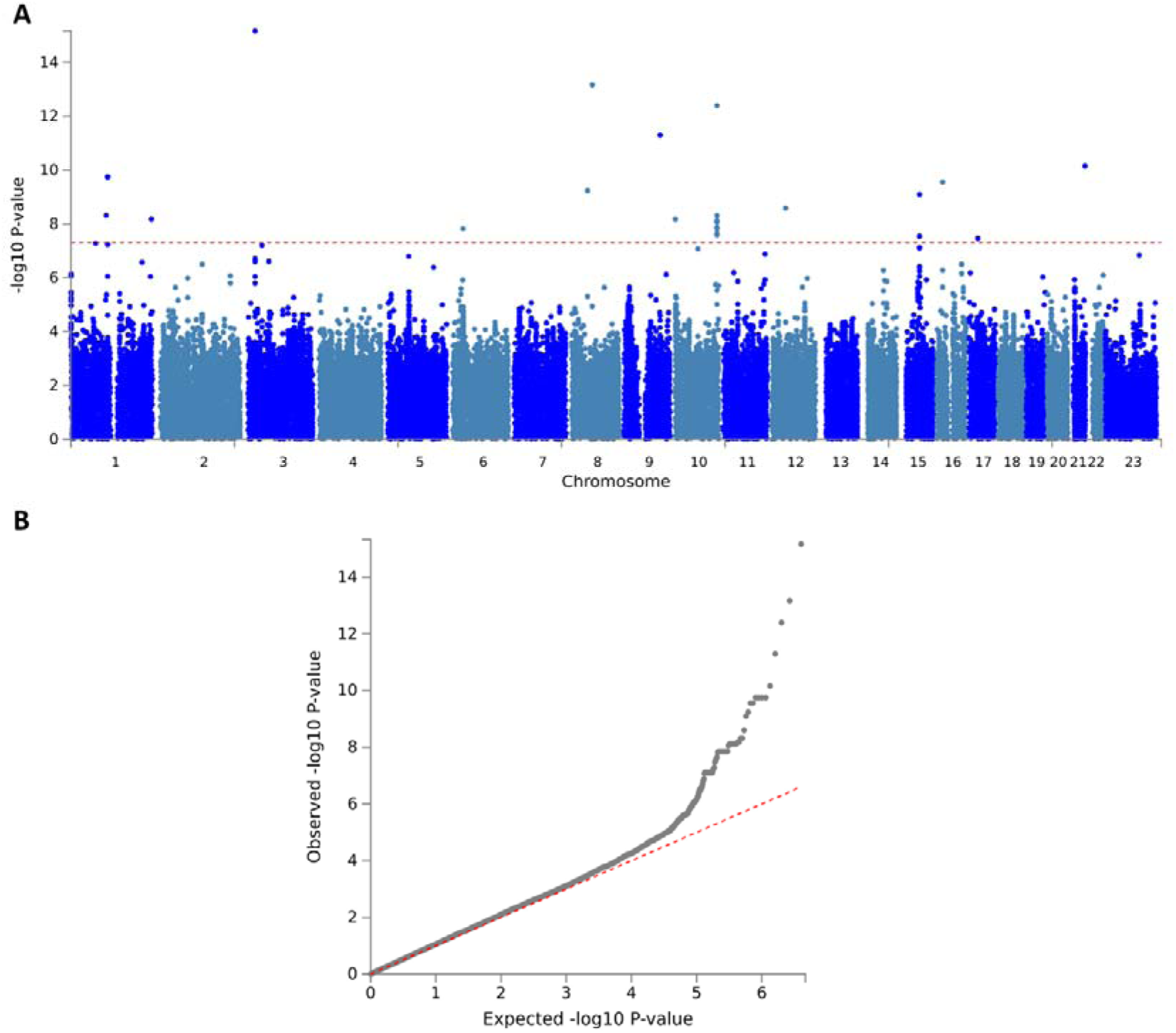
Manhattan plot and QQ-plot for migraine susceptibility. A – Manhattan plot for migraine risk in the Portuguese population. The red line indicates the *p*-value threshold for genome-wide significance (5 × 10^−8^). B – QQ-plot for migraine susceptibility in the Portuguese population.

We then assessed lead SNPs for possible regulatory functions using the RegulomeDB database. Only one significant *locus* – *RAP1A/INKA2* – likely affects TF binding and has functional evidence to affect the expression of a target gene (RegulomeDB score = 1f). eQTL data suggest that this lead SNP rs61791765 affects the expression of *RAP1A* in the thyroid tissue and *INKA2* in the esophagus tissue. Additionally, four risk *loci* (*ZNF385D*, *SMYD3*, near *ADARB2* and AC092079.1) seem to have minimal TF binding evidence (RegulomeDB score = 5, or RegulomeDB score = 6, for the AC092079.1 risk *locus*) (**Table 3**).

**Table 3.** Annotation of potential regulatory lead SNPs in RegulomeDB categories.

| Locus | SNP | RegulomeDB score | Description |
| --- | --- | --- | --- |
| *ZNF385d* | rs9812687 | 5 | TF binding or DNase peak |
| *YTHDF3* | rs4739066 | 7 | No data |
| *NPS* | rs11018195 | 7 | No data |
| *RAP1A/INKA2* | rs61791765 | 1f | eQTL+TF binding/DNase peak |
| CTA-481E9.4/ CTA-481E9.3 | rs4781860 | 7 | No data |
| AC092079.1 | rs9806148 | 6 | Motif hit |
| *PPHLN1* | rs117423705 | 7 | No data |
| *SMYD3* | rs1934582 | 5 | TF binding or DNase peak |
| AL355597.1 (near *ADARB2*) | rs118096186 | 5 | TF binding or DNase peak |

eQTL – expression quantitative trait *loci*; TF – transcription factor.

According to HaploReg eQTL data, the SNP rs4739066 (*YTHDF3*) possibly affects the expression of GGH in whole blood. Detailed information about the potential regulatory lead and candidate SNPs is shown in supplementary material, including HaploReg analysis (**Supplementary Table 2**).

### Enrichment analysis

We identified 29 GO terms significantly associated with six risk *loci* (*p* < 0.05) (**Supplementary Table 3**). Highlighted in blue are GO terms for *NPS*, *ADARB2*, *RAP1A* and *YTHDF3* genes which relate to known or hypothesized mechanisms of migraine pathophysiology **(Fig 2).** No statistically significant pathways were identified in Reactome and KEGG.

**Figure 2:**
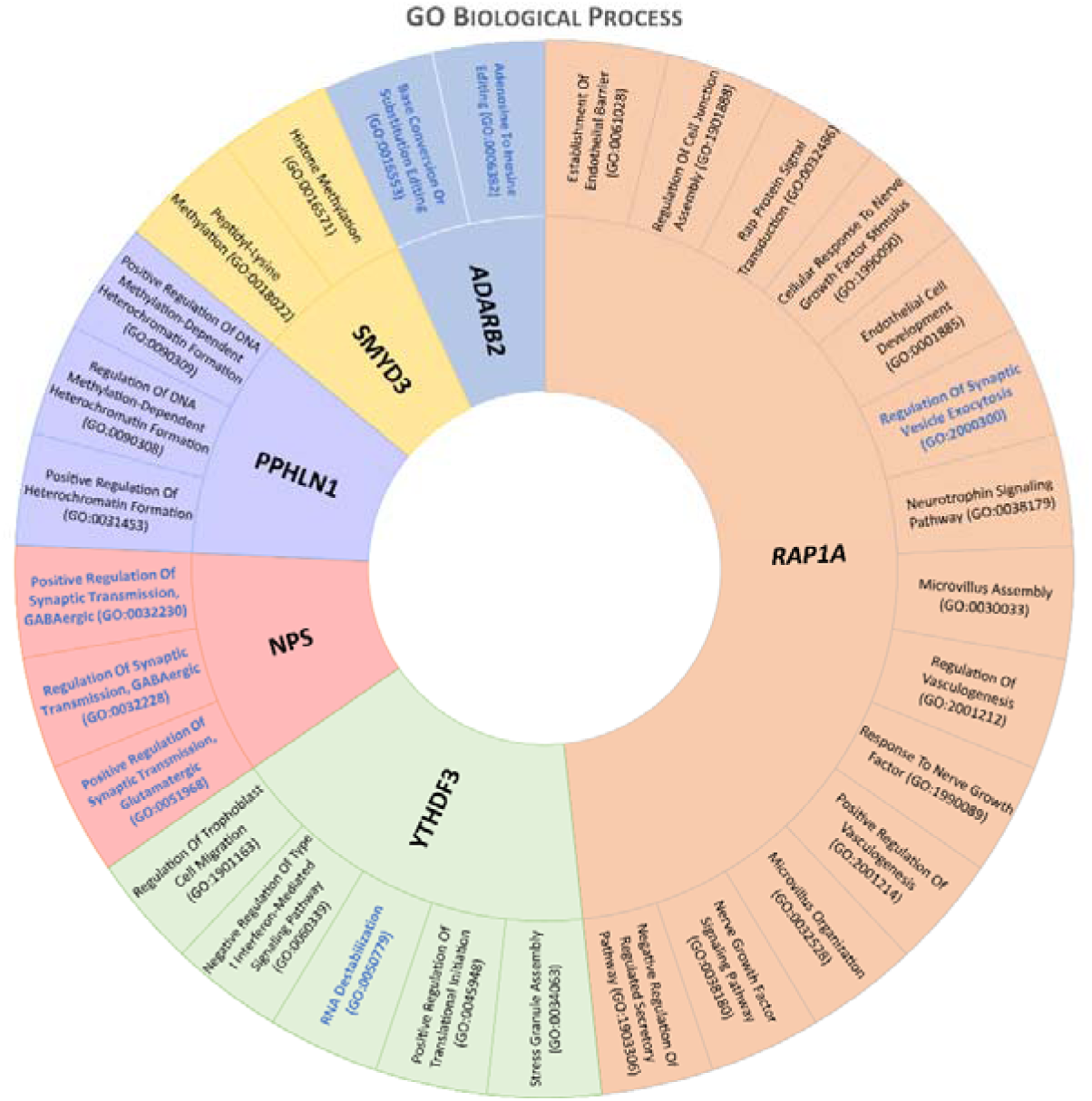
A diagram containing the statistically significant (*p*-value < 0.05) GO terms for the 6 risk *loci.* Highlighted in blue are GO terms for *NPS*, *ADARB2*, *RAP1A* and *YTHDF3* genes which relate to known or hypothesized mechanisms of migraine pathophysiology.

### Four new risk *loci* potentially associated with migraine susceptibility

Four of the new risk *loci* contain genes (*YTHDF3*, *NPS*, *RAP1A/INKA2* and near *ADARB2*) that codify proteins closely related to targets of migraine pathways. We observe a significant association at chromosomes 8 and 10 that contain the *YTHDF3* gene (lead SNP rs4739066, *p* = 6.83 × 10^−14^; **Fig 3A**) and the *NPS* gene (lead SNP rs11018195, *p* = 4.04 × 10^−13^; **Fig 3B**), respectively. Additionally, we found another *locus* on chromosome 1 containing *RAP1A/INKA2* genes (lead SNP rs61791765, *p* = 1.79 × 10^−10^; **Fig 3C**) and a *locus* on chromosome 10 near the *ADARB2* gene (lead SNP rs118096186, *p* = 6.75 × 10^−9^; **Fig 3D**).

**Figure 3:**
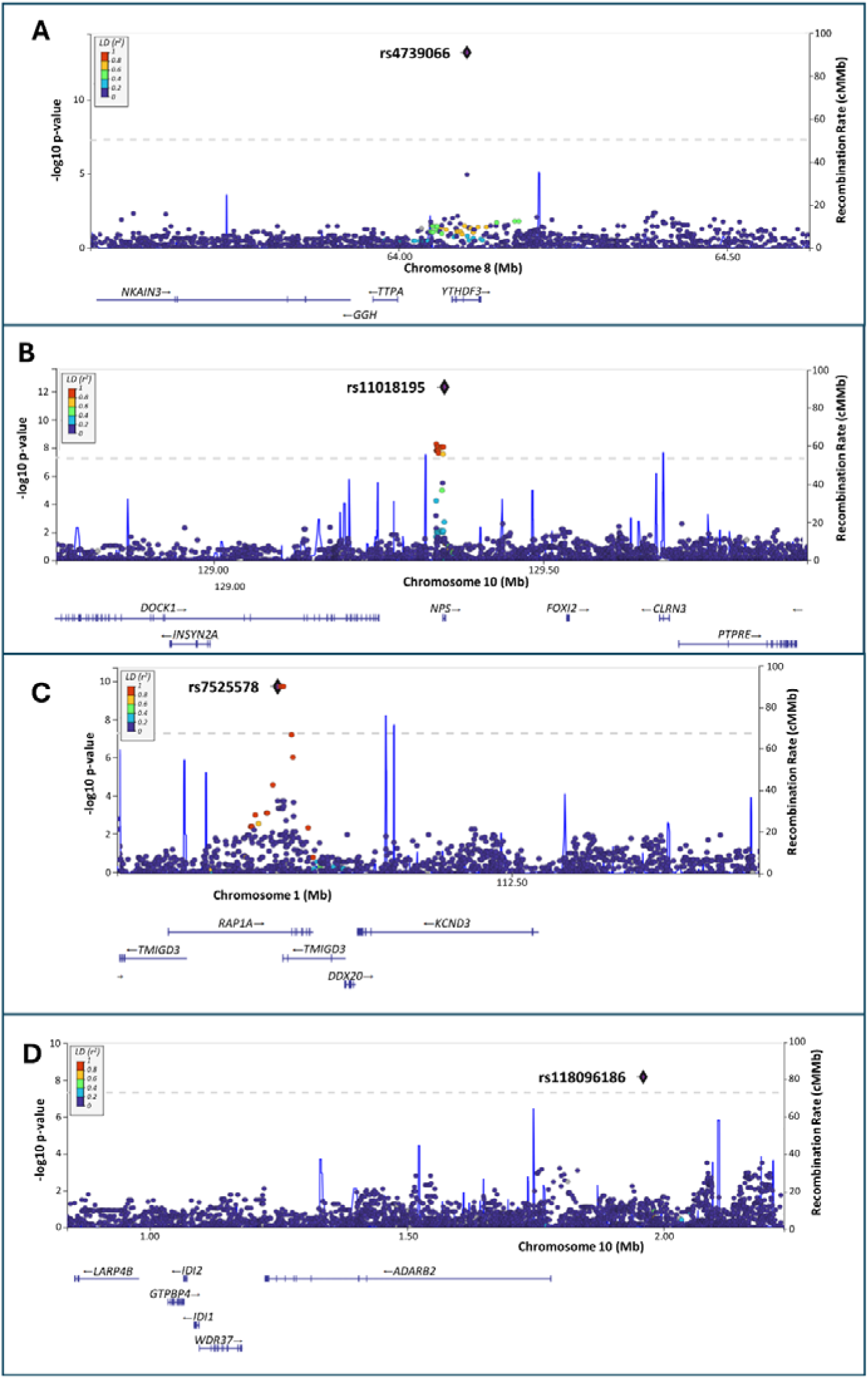
*Locus* zoom plots of four genes potentially associated with migraine pathways. A – The top associated SNP [rs4739066] for *YTHDF3* gene; B – *Locus* containing *NPS* gene, with the top associated SNP [rs11018195]; C – *Locus* containing *RAP1A* and *INKA2* genes with the top associated SNP [rs61791765]; D – Top associated SNP [rs118096186] near *ADARB2* gene. Lead SNPs are shown as a purple circle encased in a black diamond, and the remaining SNPs with colors indicate the level of linkage disequilibrium (r^2^) with the top lead SNP. The x-axis shows the chromosomal location, and the y-axis shows the uncorrected two-sided negative log10 of the *p*-value from the inverse–variance weighted fixed-effects meta-analysis. Horizontal line corresponds *to p* = 5 × 10^−8^. The underlying blue graph shows the recombination rate. Genes present at each given *locus* are displayed under their corresponding zoom plot.

Enabled by the STRING database, a rather interesting and cohesive network of proteins, backed by literature and curated databases, became apparent for the protein encoded by *NPS*, strikingly composed of proteins already known or suspected to be involved in migraine pathways (**Fig 4**). In fact, neuropeptide S (NPS) establishes interactions with several proteins encoded by genes closely linked to migraine therapies, such as ligands, receptors and transporters associated to CGRP (represented by the CALC and RAMP protein families), amylin (IAPP) and PACAP (ADCYAP1 and VIPR1) (**Fig 4**). We also performed the interactions analysis for the other three *loci* whose results are found in supplementary material (**Fig S1**).

**Figure 4:**
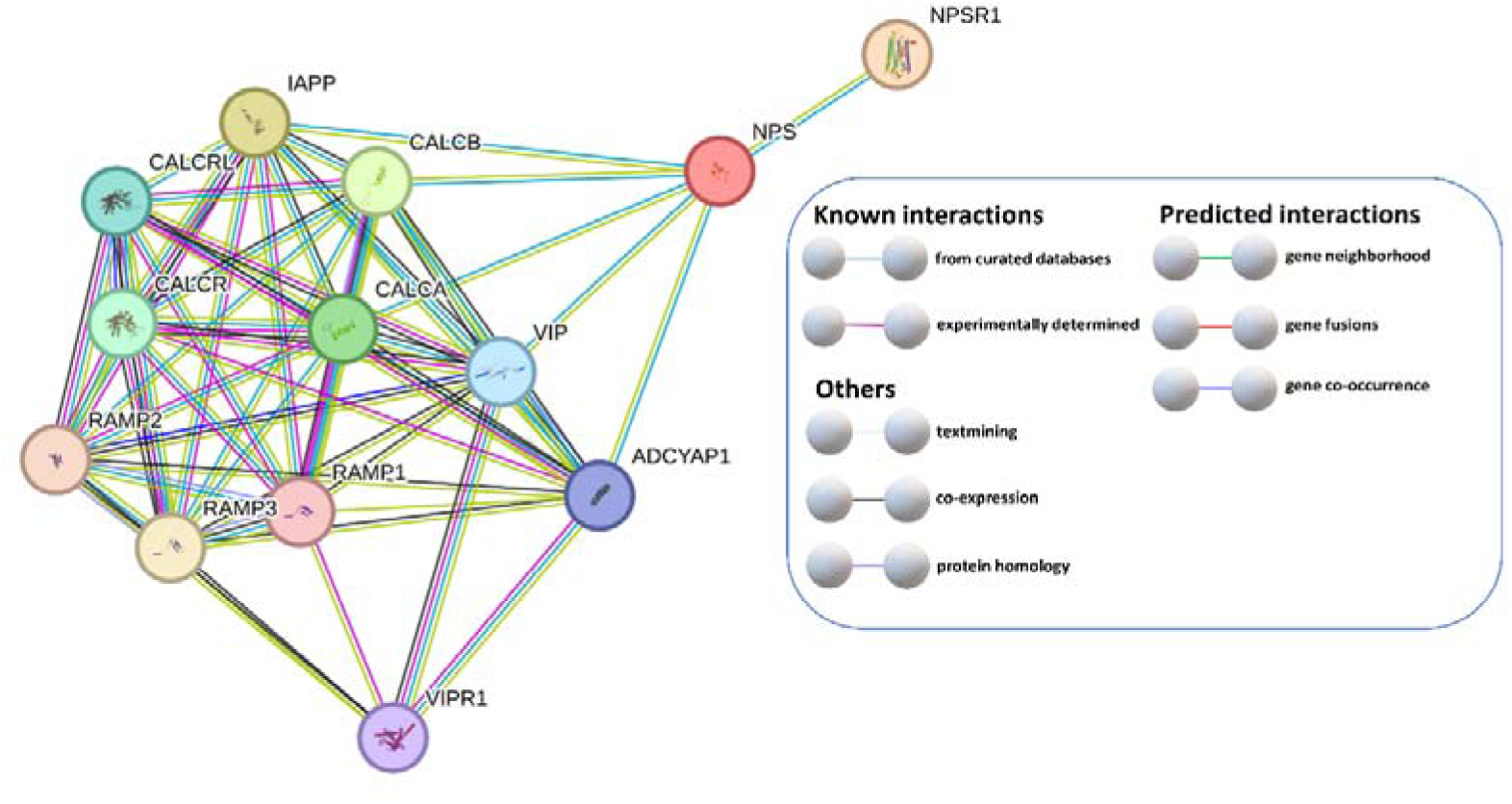
NPS establishes interactions with several proteins encoded by genes strongly associated with migraine therapies. Protein interaction map generated using the STRING database (minimum required interaction score set at 0.400). Colored lines between the proteins indicate the various types of interaction evidence.

### Polygenic risk score

We report increases of pseudo-R² following decreases of the *p*-value limit, meaning the best predictive models are, purportedly, the ones with a tighter threshold (**Fig S2**). However, this may reflect an overfitting effect – in order to avoid exacerbated overfitting while maintaining a reasonable pseudo-R^2^ of 0.26, we considered a model with a threshold of *p* = 5×10⁻⁵ to be appropriate to apply to our genotyping data.

We analyzed the 1000 SNPs with the lowest *p*-values to determine whether any previously reported migraine-associated variants were present. This analysis was automated and based on the findings reported in reference ^28^. Eleven promising variants were identified (summarized in **Table 4**), none of them having previously been associated with migraine, and are implicated in interesting pathways and possible pathophysiological mechanisms. The variants rs7690171 (*RASGEF1B*, *p* = 1.39 × 10^-8^) and rs7525578 (*RAP1A*, *p* = 1.39 × 10^-8^), both located in intronic regions, appear to be the most promising variants to migraine susceptibility. HaploReg analysis showed the latter variant is in LD (r^2^ = 1) with the lead SNP found in the GWAS for the same gene (rs61791765, *RAP1A*/*INKA2*), further supporting this hypothesis (**Supplementary Table 2**).

**Table 4.** Top SNPs with the highest significance (least p-value).

| SNP Id | P | Effect size | Location (GRCh38) | Clinical Significance | Functional consequence |
| --- | --- | --- | --- | --- | --- |
| rs117797734 | $3.31 \times 10^{-11}$ | 1.706 | 14:97,817,989 | VUS | Intergenic variant |
| rs75485498 | $5.06 \times 10^{-11}$ | 1.714 | 1:145,740,566 | VUS | RNF115 (3'UTR)<br>ENST00000582693 |
| rs7690171 | $1.08 \times 10^{-10}$ | 1.669 | 4:81,449,191 | VUS | **RASGEF1B-**<br>**intronic**<br>**ENST00000264400** |
| rs76649197 | $7.22 \times 10^{-10}$ | 1.536 | 16:18,067,009 | VUS | lncRNA<br>ENST00000563866 |
| rs7525578 | $1.39 \times 10^{-8}$ | 1.377 | 1:111,674,338 | VUS | **RAP1A-Intronic**<br>**ENST00000356415** |
| rs10815566 | $1.07 \times 10^{-5}$ | -1.093 | 9:7,239,414 | VUS | Intergenic variant |
| rs6714816 | $2.23 \times 10^{-5}$ | -0.943 | 2:126,618,590 | VUS | Intergenic variant |
| rs4964156 | $3.17 \times 10^{-5}$ | 0.880 | 2:105,731,003 | VUS | CASC18-lncRNA<br>ENST00000547465 |
| rs6562972 | $3.2 \times 10^{-5}$ | -1.061 | 13:76,856,348 | VUS | Regulatory variant<br>ENST00000377474 |
| rs62263931 | $3.89 \times 10^{-5}$ | 1.538 | 3:122,704,403 | VUS | PARP14-Intronic<br>ENST00000460683 |
| rs7839555 | $4.25 \times 10^{-5}$ | 1.060 | 8:19,397,488 | VUS | Intergenic variant |

Variants ordered by increasing *p*-values (P), accompanied by chromosome and genomic location, clinical significance and functional consequence. RAP-related variants highlighted in bold.

## DISCUSSION

This study reports, for the first time in the Portuguese population, a pilot GWAS to uncover genetic factors that could be involved in migraine susceptibility and followed up with the calculation of PRS. GWAS identified a total of nine *loci* and, among these, four *loci* (*YTHDF3*, *NPS*, *RAP1A*, near *ADARB2*) seem to be involved in well-established pathophysiological pathways of migraine. The mechanistic relevance of other identified *loci* still needs to be ascertained. Regarding the PRS analysis, the models’ predictive capabilities were directly linked to the stringency of the *p*-value thresholding; having selected a reasonable model, it is noteworthy that none of the most significant variants had any previous indication of pathogenicity in the context of migraine disorders.

It is important to underscore that the limited replication of GWAS across different populations can be attributed to variations in genetic architecture. Ethnically diverse groups exhibit differences due to population-specific variations and changes in allele frequency resulting from genetic drift, local selection, or both. Moreover, founder effects are known to play a role in prevalence variations of complex diseases among populations ^8^. Cross-study comparisons indicate potential variations in migraine prevalence based on ethnicities, which could stem from methodological aspects, ethnicity-specific variations in genetic predisposition, environmental risks, or cultural influences affecting symptom reporting ^29^. These differences between populations can translate into different clinical manifestations and disparities in therapeutic response. Often, diverse populations are studied as part of large meta-analyses that combine data to estimate associations to identify variants with consistent effects across populations, but this hinders the detection of population-specific genetic risk factors ^8^.

### Three risk *loci* are associated with other neurological disorders

Three of the identified *loci* (*ZNF385D*, *PPHLN1* and *SMYD3*) were already associated with other neurological diseases, but evidence of their association with migraine is still limited.

The most significant lead SNP is located within the *ZNF385D* gene and has been associated with neuropsychiatric disorders, such as schizophrenia ^30^, reading disability and language impairment ^31^, and epilepsy ^32^ – pathologies associated with migraine comorbidities. An enrichment analysis reported numerous connections between epilepsy and migraine regarding ion channels, regulation of membrane potential, and transmembrane transport. Both pathologies are linked to abnormal neuronal excitation, suggesting that cortical spreading depression plays a shared role in the development of these disorders. Owing to the multitude of molecular mechanisms shared between epilepsy and migraine, various antiepileptic drugs have been investigated and endorsed for migraine prevention ^33^.

The ninth top lead SNP is located within the *PPHLN1* gene, which encodes periphilin-1, an important protein in the nervous system that has been shown to play a crucial role in early embryogenesis in mice. Research has implicated periphilin-1 in the etiology of Parkinson’s disease, suggesting its potential relevance to this neurological condition ^34^. Moreover, recent studies have revealed that *PPHLN1* is involved in mediating epigenetic silencing ^35^. Indeed, epigenetics has come to offer a promising avenue for delving deeper into the explanation of migraine development ^36^.

The eleventh top lead SNP is located within the *SMYD3* gene, which influences transcriptional regulation and the methylation of various histone and non-histone targets, contributing to cancer regulation. Studies have indicated potential associations of the *SMYD3* with intellectual disability, infertility, and autism ^37^. Additionally, SMYD3 activity has significant connections to the immune system and blood cancers ^38^. It is noteworthy that pre-clinical data continues to substantiate the role of pro-inflammatory cytokines and immune cells in both initiation and maintenance of headaches ^39^.

### *YTHDF3* and *ADARB2* genes are involved in RNA-editing machinery

Two of the statistically significant *loci* are associated with genes (*YTHDF3* and *ADARB2*) involved in RNA-editing mechanisms. RNA-editing plays an important role in regulating synaptic function and can occur through an adenosine-to-inosine (A-to-I) edit. Disrupted A-to-I RNA-editing has been associated with various neurological and neurodegenerative disorders, such as amyotrophic lateral sclerosis, Alzheimer’s disease, Huntington’s disease, epilepsy, depression, and schizophrenia ^40^. A-to-I conversion through hydrolytic deamination is catalyzed by adenosine deaminases acting on RNA (ADARs) – this editing process alters the RNA sequence and has been identified in both protein-coding and non-coding transcripts of neuronal cells ^40^. Intriguingly, methyl modifications in mRNAs of synaptic proteins have also been identified, suggesting an involvement of mRNA modifications in synaptic plasticity ^41^. In fact, the internal modification of eukaryotic mRNA known as N6-methyladenosine (m6A) plays a crucial role in regulating gene expression and mRNA stability ^42^. Particularly, the presence of m6A can inhibit ADAR binding and decrease A-to-I editing of specific targets ^41^.

#### YTHDF3

YTH domain-containing proteins (YTHDF1-3) recognize methylated RNA. Interestingly, while generally binding to the same mRNAs, they exert distinct effects on their targets. All of them are found in dendrites and colocalize with synaptic markers, suggesting their potential involvement in local regulation of synaptic mRNAs ^41^. Furthermore, all three proteins have been observed in neuronal RNA granules, highlighting potential roles in mRNA transport ^42^.

YTHDF3 is a versatile player in both the translational efficiency and degradation of m6A-modified mRNA – a dual role in determining the fate of m6A-modified mRNA that warrants further investigation ^43^. In large-scale studies, likely disruptive *YTHDF3 de novo* variants were found to be significantly enriched in individuals with intellectual disability and/or developmental delay ^44^. YTHDF3 knockdown using siRNA oligonucleotides has been shown to reduce the translation efficiency and decay of m6A-modified mRNA. Additionally, m6A methylation plays a critical role in both the regulation of cardiac disease and the development of arterial disease, with implications in abdominal aortic aneurysm (AAA) incidence and rupture ^42, 45^. Strikingly, patients with higher YTHDF3 expression were also at greater risk of AAA rupture ^46^, and a separate study observed that YTHDF3 expression levels were significantly different between myocardial infarction (MI) tissue samples and those of non-failing controls ^47^.

An increasing number of studies have investigated the association between migraine and vascular disease. In that regard, a study conducted by Daghlas *et al*., reported a genome-wide genetic correlation between CeAD (cervical artery dissection) and migraine, particularly MwoA. The findings emphasize a shared genetic risk between migraine and CeAD while identifying *loci* with likely vascular functions in migraine ^48^. Furthermore, a GWAS demonstrated a weak association between the *YTHDF3* rs4739066 variant (the lead SNP identified in this study) and MI in the Saudi Arabian population ^49^. More recently, a meta-analysis demonstrated that migraine was associated with a greater risk of MI, and ischemic, hemorrhagic and unspecified strokes ^50^. These findings point to a role of m6A methylation and its regulatory factors in influencing inflammation within the vascular system. In this GWAS, we found that the lead SNP of *YTHDF3* acts as an eQTL by affecting the expression of *GGH* in whole blood – a gene encoding an enzyme with a role in facilitating the export of cellular folate. Previous studies have identified polymorphisms in the *GGH* region that are linked to heightened promoter activity and increased DNA uracil content. Moreover, the *GGH* gene has been linked to reduced cardiovascular disease (CVD) risk, and this association is particularly prominent in cases of early onset CVD ^51^.

#### ADARB2

*ADARB2* encodes a catalytically inactive protein primarily expressed in the brain, amygdala, and thalamus. Despite its catalytic inactivity, ADARB2 has been found to act as a competitor of ADAR and ADARB1 *in vitro*. By binding to the same transcripts, ADARB2 can reduce the efficiency of other RNA-editing enzymes, preventing substrate RNA deamination in brain regions where they are co-expressed. Consequently, ADARB2 may decrease the overall number of edited transcripts through this competitive mechanism, potentially leading to impaired conductance and synaptic dysfunction in the central nervous system ^52^.

In 2012, Griffiths *et al*. performed a pedigree-based GWAS of the Norfolk Island population and reported four *ADARB2* SNPs that were positively associated with migraine susceptibility ^53^. Later, they decided to investigate SNPs in *ADARB2*, as well as in *ADARB1*, in a case-control of Australian population. However, this study did not find any evidence supporting the association between the two RNA-editing genes and an increased risk of migraine – possibly because genetically isolated populations can possess unique genetic characteristics that may not always be replicated in broader case-control cohorts ^52^.

### Ras-family small GTPases involved in synapse plasticity and immune response

Ras-family small GTPases are a class of guanine-nucleotide-binding proteins that function as molecular switches, transitioning between active (GTP-bound) and inactive (GDP-bound) states. Their activation is regulated by guanine nucleotide exchange factors (GEFs), which mediate replacement of GDP with GTP, and by GTPase-activating proteins (GAPs), which increase the intrinsic hydrolysis reaction of GTP to GDP by several orders of magnitude ^54^.

Within this family, RAP1 is a highly conserved 21-kDa monomeric small GTPase, expressed in two isoforms – RAP1A and RAP1B ^55^. Human RAP1 proteins are closely related to RAP2 subfamily members, and some identical or overlapping functions have been described for RAP1 and RAP2 proteins ^54,56^. In neuronal cells, specifically, RAS, RAP1 and RAP2 play distinct roles and are selectively activated by various forms of synaptic activity. This activation occurs through the stimulation of NMDA receptors and the subsequent influx of calcium. Importantly, these GTPases independently regulate three different activity-dependent trafficking events of AMPA receptors ^57^. RAS and RAP1 have been proposed to mediate the stimulatory effects of certain G protein-coupled receptors on extracellular signal-regulated kinase (ERK) activity. In line with this, a previous study demonstrated that PACAP, which activates a G protein-coupled receptor to induce neuronal differentiation via ERK1/2 activation, transiently activates RAS and induces sustained GTP loading of RAP1, thus enhancing GTP loading of both RAP1 and RAS. RAP1 activation was found to be crucial for PACAP-induced ERK stimulation together with the activity of protein kinase A (PKA), protein kinase C (PKC), and calcium-dependent signaling pathways ^58^. As aforementioned, PACAP is an important neurotransmitter in migraine pathophysiology. Thus, the success of anti-CGRP therapies can possibly be reproduced with the use of monoclonal antibodies targeting PACAP ^59^.

Ras-GEF domain-containing member 1b (RASGEF1b) belongs to a small protein subgroup whose members – RASGEF1a, RASGEF1b, and RASGEF1c – share the conserved REM and catalytic CDC25-HD domains, but lack additional common structural features. RASGEF1b displays specific GEF activity toward the RAP2 GTPase, while showing no detectable activity toward RAP1 or other members of the Ras subfamily. RASGEF1b plays important roles in immune cells, particularly in macrophages, with RASGEF1b-deficiency leading to decreased gene expression under resting conditions, potentially altering homeostatic signaling and affecting responsiveness to danger signals. RASGEF1b activates RAP2A by catalyzing its transition from a GDP-bound inactive state to a GTP-bound active form, thereby initiating downstream signaling pathways that regulate multiple cellular processes. RAP2A expression in macrophages appears to be tightly controlled, partly through NF-κB, to maintain balanced immune responses ^60^.

### Neuropeptide S and its receptor may act as a key synaptic neurotransmitter system

A risk *locus* uncovered in our GWAS pertains to the *NPS* gene, whose protein NPS exhibits selective binding and activation of the neuropeptide S receptor 1 (NPSR1, or GPR154). NPSR1 belongs to the G protein-coupled receptor family, and the combination of NPS/NPSR1 acts as a neurotransmitter system in the brain. Activation of NPSR1 by NPS leads to intracellular signaling events, including calcium mobilization, increase of cyclic adenosine monophosphate (cAMP) levels, and activation of the MAPK pathway ^61,62^. NPS and NPSR1 are predominantly expressed in specific regions of the brain, such as the amygdala, hypothalamic nucleus, and hippocampus.

A previous study has revealed additional effects of NPS, namely as transcriptional upregulation of proinflammatory cytokines, such as interleukin 8 and substance P, upon transfection with NPSR1 in epithelial cells ^62^. Another study conducted by Jüngling *et al*. demonstrated that NPSR1 activation enhances glutamatergic transmission from principal neurons in the lateral amygdala to GABAergic paracapsular intercalated cells. This suggests that NPSR1-mediated calcium mobilization promotes an increase in neurotransmitter release, leading to enhanced synaptic transmission ^63^. Strikingly, NPS interacts with the isoforms of CGRP, encoded by *CALCA* and *CALCB*, and amylin (IAPP). This direct relation to genes and pathways connected to migraine and migraine-specific therapeutics creates a promising rationale for further studies on *NPS*.

### Prediction of treatment response

Although patient medication response was not directly assessed, the PRS approach can distinguish genetic variants associated with treatment outcomes in migraine. Using GWAS data from 375,000 individuals, a PRS model evaluated response to common acute migraine therapies. Despite limited effect sizes and lack of clinical applicability, the PRS model successfully explained patient response to triptans, supporting the potential of precision medicine in migraine ^64^.

Our study is mainly limited by the relatively small sample size. Moreover, it builds upon a cohort of patients that stem from a single tertiary hospital, receiving referrals of complex cases, and thus preventing generalizations into community settings. Furthermore, our data is population-specific and thus findings cannot be extrapolated and applied to the global population.

According to established guidelines ^25^, *“if the GWAS data are relatively underpowered, the optimal threshold is more likely to be a P-value of 1.”* In our case, the situation is reversed, because the number of individuals in both the test and validation datasets is small relative to the number of SNPs analyzed – the model naturally tends to focus on the most relevant variants, shifting the optimal *p*-value threshold closer to 0. Nevertheless, the risk of overfitting must be carefully considered: relevant SNPs may be overlooked in favor of those showing stronger, but potentially spurious, statistical signals, thereby reducing the model’s accuracy and generalizability. Conversely, applying overly stringent *p*-value thresholds may inadvertently exclude variants that are genuinely associated with the phenotype, or which contribute to the pathophysiology of migraine ^25^.

It is also important to emphasize that a statistical association between a phenotype and a SNP does not necessarily indicate a causal relationship. A non-functional SNP may appear associated with the phenotype due to strong linkage disequilibrium with the true causal variant. If the biologically meaningful SNP is removed during quality control – owing to low MAF, Hardy-Weinberg disequilibrium, or missing data – the clumping process may instead retain a proxy SNP as the regional representative. Such substitutions can obscure the true genetic signal and complicate the biological interpretation of the results ^65^.

We report herein the first pilot migraine GWAS in the Portuguese population, detecting nine risk *loci* for migraine susceptibility. The findings reinforce the importance of neurotransmitter release and synaptic transmission, as well as the involvement of the vascular component in migraine pathophysiology. Among our results, the discovery of a new *locus* within the *NPS* gene stands out as a potential new molecular target directly related to recently developed and effective migraine treatments (CGRP and amylin therapies). This work further underscores that GWAS can provide novel, clinically valuable insights into populational and disease-associated genetic landscapes, further enabling therapeutic developments and bolstering personalized medicine approaches.

## ACKNOWLEDGEMENTS

We would like to acknowledge all patients and healthy controls for participating in this study.

## STATEMENTS AND DECLARATIONS

### Authors contributions

Conceptualization: Santos, M; Gil-Gouveia, R; Alves-Ferreira, M; Azevedo, L; Lemos, C and Dias, A. Investigation: De Marco, R; Pucci, K and Dias, A. Methodology: De Marco, R; Pucci K; Cavadas, B and Dias, A. Writing-original draft: De Marco, R; Santos, M and Dias, A. Writing-review & editing: Santos, M; Gil-Gouveia, R; Sousa, A; Alves-Ferreira, M; Azevedo, L; Lemos, C. and Dias, A. All authors have read and agreed to the published version of the manuscript.

### Statement on the use of AI tools

Supervised use of AI tools (ChatGPT, GPT-5.1) was employed to improve the language and readability of the manuscript.

### Data availability statement

The dataset supporting the conclusions of this article is included within the article and its additional files. The full data generated or analyzed during this study can be made available upon reasonable request to the corresponding author.

### Ethics approval and consent to participate

Institutional Review Board Statement: The use of biological material and information from patients was approved by the Committee for Ethical and Responsible Conduct of Research CECRI, i3S; approval code 2/CECRI/2020. Written informed consent was obtained from all subjects involved in the study.

### List of Abbreviations

AAA: abdominal aortic aneurysm
ADARs: adenosine deaminases acting on RNA
CADD: combined annotation dependent depletion
CGRP: calcitonin gene-related peptide
CVD: cardiovascular disease
eQTL: expression quantitative trait loci
ERK: extracellular signal-regulated kinase
FHM: familial hemiplegic migraine
GO: gene ontology
GWAS: genome-wide association study
HWE: Hardy-Weinberg equilibrium
KEGG: kyoto encyclopaedia of genes and genomes
LD: linkage disequilibrium
m6A: N6-methyladenosine
MAF: minor allele frequency
MI: myocardial infarction
MwA: migraine with aura
MwoA: migraine without aura
NPS: Neuropeptide S
PACAP: pituitary adenylate cyclase-activating peptide
PRS: polygenic risk score
QC: quality control
QQ: quantile-quantile
SD: standard deviation
SNP: single nucleotide polymorphism
TF: transcription factor
VIP: vasoactive intestinal peptide
VUS: variant of unknown significance

---

**Figure S1:** Protein interaction map for RAP1A/INKA2, YTHDF3 and ADARB2, generated using the STRING database (minimum required interaction score set at 0.400). Colored lines between the proteins indicate the various types of interaction evidence.

**Figure S2:** Pseudo-R² value plotted relative to the *p*-value thresholds set for the PRS model.

