## Supplementary figures and images for "*Neuropeptide S* as a potential risk *locus* for migraine in the Portuguese population"

### Figure S1

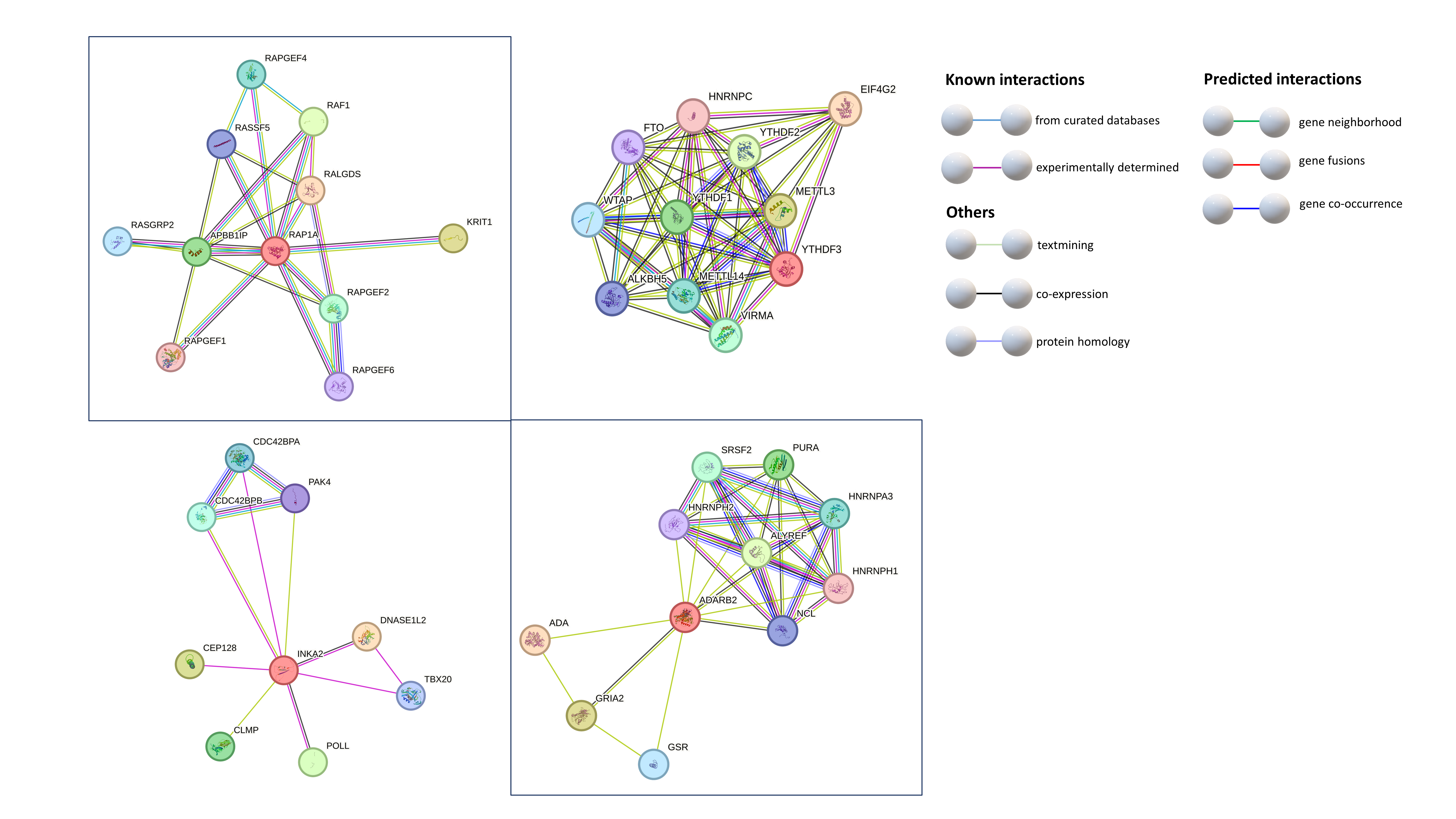

### Figure S2

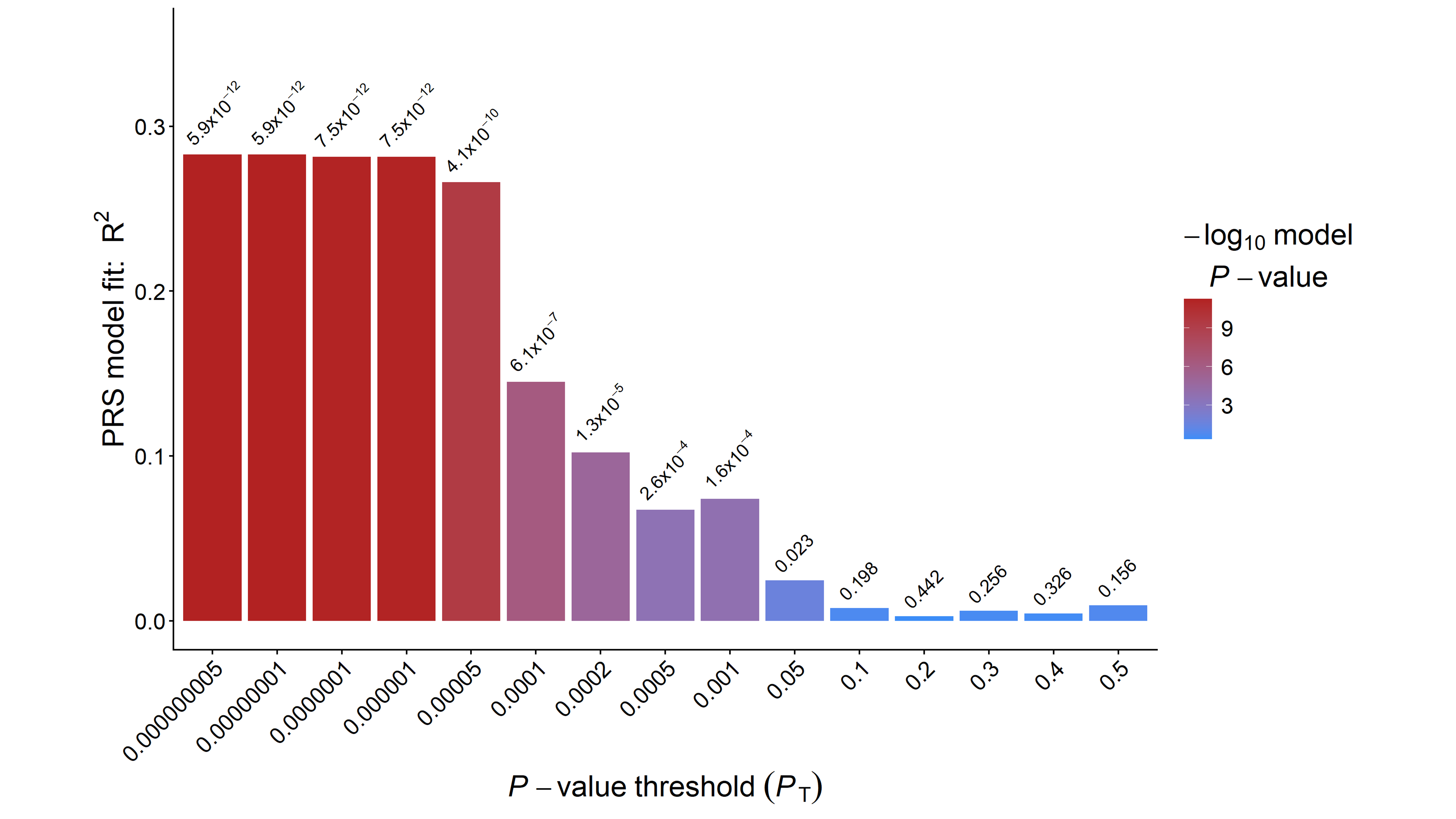
